# Impact of first SARS-CoV-2 infection variant on serological responses against Omicron: findings from the SIREN study

**DOI:** 10.1101/2025.06.02.25328497

**Authors:** Ferdinando Insalata, Ana Atti, Edward J. Carr, Ashley D. Otter, SIREN study group, Sarah Foulkes, Giulia Dowgier, Agnieszka Hobbs, Mary Y. Wu, Susan Hopkins, Andre Charlett, Rupert Beale, Victoria Hall

## Abstract

**Background:** Despite the existing hybrid immunity, a sharp increase in SARS-CoV-2 reinfections was observed worldwide following Omicron variant emergence. We investigated whether the first infecting variant indelibly shapes serological responses against Omicron (BA.1 and BA.2) reinfection.

**Methods:** Participants with a sequence-confirmed Alpha (n=23) or Delta (n=10) first infection before third vaccine dose (V3) that subsequently had a BA.1 or BA.2 reinfection were selected. Sera were tested for anti-SARS-CoV-2 spike (anti-S) and live virus microneutralisation (LV-N) against Ancestral, Alpha, Delta, Omicron BA.1 and BA.2. Antibody responses and waning post-V3 were compared by first infection variant using mixed-effect models, as well as inferred titres 7-days before reinfections. Individual’s neutralisation responses were compared 12 weeks post-V3, among those with Alpha and Delta primary infection.

**Results:** After V3, those with Delta first infection had higher LV-N Omicron BA.1 titres (fold difference (FD)=2.7, p=0.05) compared to Alpha primary infection. Participants with Delta first infection presented higher LV-N BA.1 (FD=1.89, p=0.004) and LV-N BA.2 (FD 2.06, p=0.001) titres pre-Omicron reinfections. Individuals’ neutralisation responses against Ancestral were higher than any other subsequent variants, regardless of first infection variant.

**Discussion:** A previous Delta SARS-CoV-2 infection induced a higher serological response against a subsequent Omicron infection when compared to Alpha first infections.

## INTRODUCTION

SARS-CoV-2 reinfections have been reported since 2020 and were considered a relatively rare event until the emergence of Omicron variants from November 2021, particularly BA.1 and BA.2 subvariants. Omicron had strong immune evasion potential due to a larger number of mutations in spike (S) compared to previous SARS-CoV-2 variants and was responsible for a remarkable increase in SARS-CoV-2 infection rates worldwide, despite most of the population being previously vaccinated, infected or both. (1–4) As expected, the first generation of vaccines showed reduced effectiveness against Omicron. As reinfection became common, it became important to understand the nature and extent of protection offered by hybrid immunity conferred by both infection and vaccination. In particular it is unclear to what extent the variant that an individual with hybrid immunity was first exposed to contributed to protection at the time of Omicron exposure. (1, 2)

During the early stages of the pandemic, previous infection was associated with high protection against SARS-CoV-2 reinfection. (5) This persisted despite the rapid spread and rise in infection rates with the emergence of the more transmissible Alpha variant in the UK (December 2020). (6, 7) Subsequently, the Delta variant has shown to be even more transmissible than Alpha and rapidly become dominant in the UK by May 2021, although vaccination reduced the transmissibility of both variants (8). A modest reduction in vaccine effectiveness was observed against Delta variant compared to Alpha after two vaccine doses. (9)

COVID-19 vaccines, although effective in preventing severe disease for long time periods, offer only short-term immunity against infection. This is due to the mismatch between vaccine antigen and emerging variants, and also due to waning in neutralising antibody titres after vaccination. (10) More substantial and longer-lasting protection has been observed in those who have been both vaccinated and infected, compared to those vaccinated without previous infection. This is presumably due to greater antigen dose and longer expression. (11–12)

In this context, a key aspect to be considered is how the exposure to viral antigens may shape the subsequent protection to related strains – also known as immunological imprinting. (13) For SARS-CoV-2, this phenomenon can be induced both by infection or vaccination and, in some interpretations of imprinting, may interfere with developing a broader immune response against emerging variants. (14) It has been reported that individuals infected with a newer SARS-CoV-2 variant who were previously exposed to Ancestral SARS-CoV-2 antigens (deriving both from infection and/or vaccination) had higher neutralisation response against Ancestral than the newer infection variant, which is sustained over time. (15–19). This could potentially explain the lower protection offered by vaccines against Omicron variants.

Within the SARS-CoV-2 Immunity and Reinfection Evaluation (SIREN) study, we aim to investigate how first SARS-CoV-2 infection variant shapes the serological response against Omicron (BA.1 and BA.2) reinfection.

## METHODS

### Study population and design

We conducted an analysis of sequential sera samples from a subset of SIREN participants, who regularly underwent SARS-CoV-2 PCR and serology testing (ISRCTN 11041050). The SIREN study protocol is described elsewhere. (20)

### Case and sample selection

We selected participants according to the criteria below:

#### Inclusion criteria

- Had a sequence-confirmed first infection by Alpha or Delta variant;
- Had a subsequent Omicron BA.1 or BA.2 infection;
- Had received a third vaccine dose (V3) before their Omicron reinfection;
- Had at least one serum sample available between V3 and their Omicron reinfection.

#### Exclusion criteria

- Had received a fourth vaccine dose before their Omicron reinfection;
- Had positive anti-SARS-CoV-2 nucleocapsid (anti-N) before their first infection (likely previously infected).

### Sample testing

All sera samples available from the selected participants were tested for anti-N antibodies, anti-SARS-CoV-2 spike (anti-S) antibodies and live virus microneutralisation (LV-N) against Ancestral, Alpha, Delta, BA.1 and BA.2 variants. Details on antibody testing is provided elsewhere (21, 22). The LV-N quantitative range is 40-2560 and, for the purpose of this analysis, titres above the quantitative range were replaced with the value at the top of the quantitative range (2560). For sera whose dilution did not cross 50% inhibitory concentration, we qualitatively reported as no neutralisation, weak neutralisation or complete neutralisation, assigning values of 5, 10 and 5120 respectively for visualisation. When calculating geometric means, we moved values outside of the quantitative range to the endpoints.

### Data analysis and modelling

#### Dynamics of antibody levels and nAbs titres post-V3 by variant of first infection

We compared at peak antibody responses and antibody waning following V3 by variant of first infection (Alpha and Delta). We reported the fold difference (FD) as being the ratio of geometric means between the two groups defined by the first-infection variant. All sera samples taken from 7 days after the third vaccine dose to assess peak antibody response, consistent with previous studies, and until 7 days before reinfection were included. (22)

Trajectories (logarithmic scale) of antibody metrics were modelled using mixed-effect models, as random intercepts at participant level were supported for all antibody responses (likelihood ratio test); for anti-S, random slopes were also supported. Time since third dose and variant of first infection were included as predictors. Differences in peak values (models intercepts) were assessed using Wald test, allowing for different decay rates by variant of first infection, via an interaction term between the variant and time variable. Overall decay rates were estimated by fitting model without the interaction term. We also controlled for time since first infection, including the period between first infection and third vaccine dose, assessing its significance via Wald test. For anti-S titres, a standard linear mixed model was used; for nAb titres, mixed effect tobit model was applied to account for left and right censoring of titres falling outside the quantitative range of the assay.

#### Comparing inferred antibody response at time of BA.1 infection by variant of first infection

We compared inferred antibody responses by variant of first infection at the time of BA.1 infections. Fitted models were used to predict the antibody levels and nAb titres at this specific time. All participants were included in this analysis. For people who were infected with BA.1 we inferred at 7 days before the PCR confirming reinfection. For people who were not infected with BA.1 (but later with BA.2) we matched time since third dose: we predicted levels/titres at 12 weeks after third dose, 7 days before the median time of a BA.1 infection (which is 13 weeks after third dose). On these inferred values, we assessed differences by variant of first infection by using *t*-test with unequal variances (Welch’s test), as the spread of data points in the groups compared was visibly different in most cases. We replaced values outside the quantitative range (full neutralisation) with the upper end of the quantitative range. On the same inferred levels and titres, we assessed differences by sub-variant of reinfection, using the same statistical test.

#### Comparing inferred antibody response immediately before BA.2 infection by variant of first infection

We compared predicted antibody responses by variant of first infection, immediately before BA.2 reinfection. Only BA.2-infected participants were included in this analysis to have comparable decay times since V3. Antibody levels and nAb titres of participants with BA.2 infection were inferred at 7 days before reinfection. We tested for differences in geometric means by variant of first infection across anti-S and different nAb titres using Welch’s test.

#### Comparing neutralisation responses among those with Alpha or Delta first infection

We compared inferred individual’s neutralisation responses against LV-N Ancestral, LV-N Alpha, LV-Delta, LV-N Omicron BA.1 and LV-N Omicron among those who had Alpha first infection or Delta first infection, at 12 weeks after V3 (proxy time of BA.1 infections). We reported the fold change (FC) as being the ratio of the values of different responses (for example, LV-N Alpha to LV-N Delta) in a participant. All participants were included in this analysis. We used the same method to infer titres at this specific time as described above. We calculated geometric means of LV-N Ancestral, LV-Alpha, LV-Delta, LV-N Omicron BA.1 and LV-N Omicron inferred titres.

## RESULTS

A total of 87 samples from 33 participants were selected from the SIREN database and fit our selection criteria. Samples from these participants taken between 08^th^ October 2021 and 07^th^ April 2022 were included in the analysis.

Selected individuals were predominantly female (82%) and of white ethnicity (88%), with a median age of 44 years (34, 52) (Table 1). First infections occurred between the 29^th^ December 2020 and 05^th^ November 2021; 70% (23/33) were Alpha and 30% (10/33) were Delta. A comparison on demographics between participants with Alpha and Delta first infection is reported in Supplementary Table 1. Patterns of antigenic exposures from all participants are described in Figure 1.

**Figure 1.**
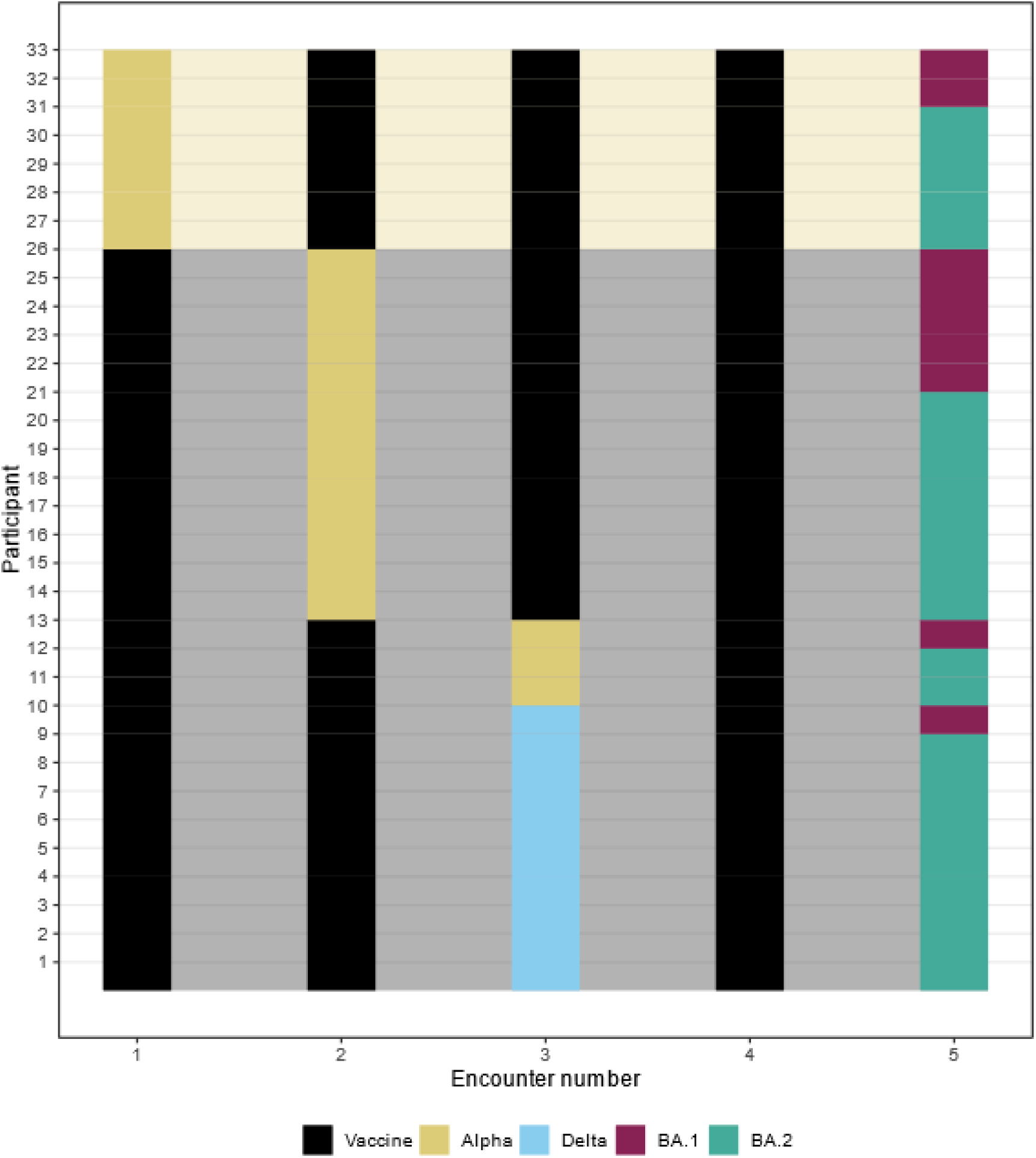
Antigenic encounters for each participant. Each line represents one participant included in this analysis. For example, Participant 13 had their first vaccine dose, followed by an Alpha first infection; after that, they had their second and third vaccine dose, followed by Omicron BA.2 infection.

**Table 1:**
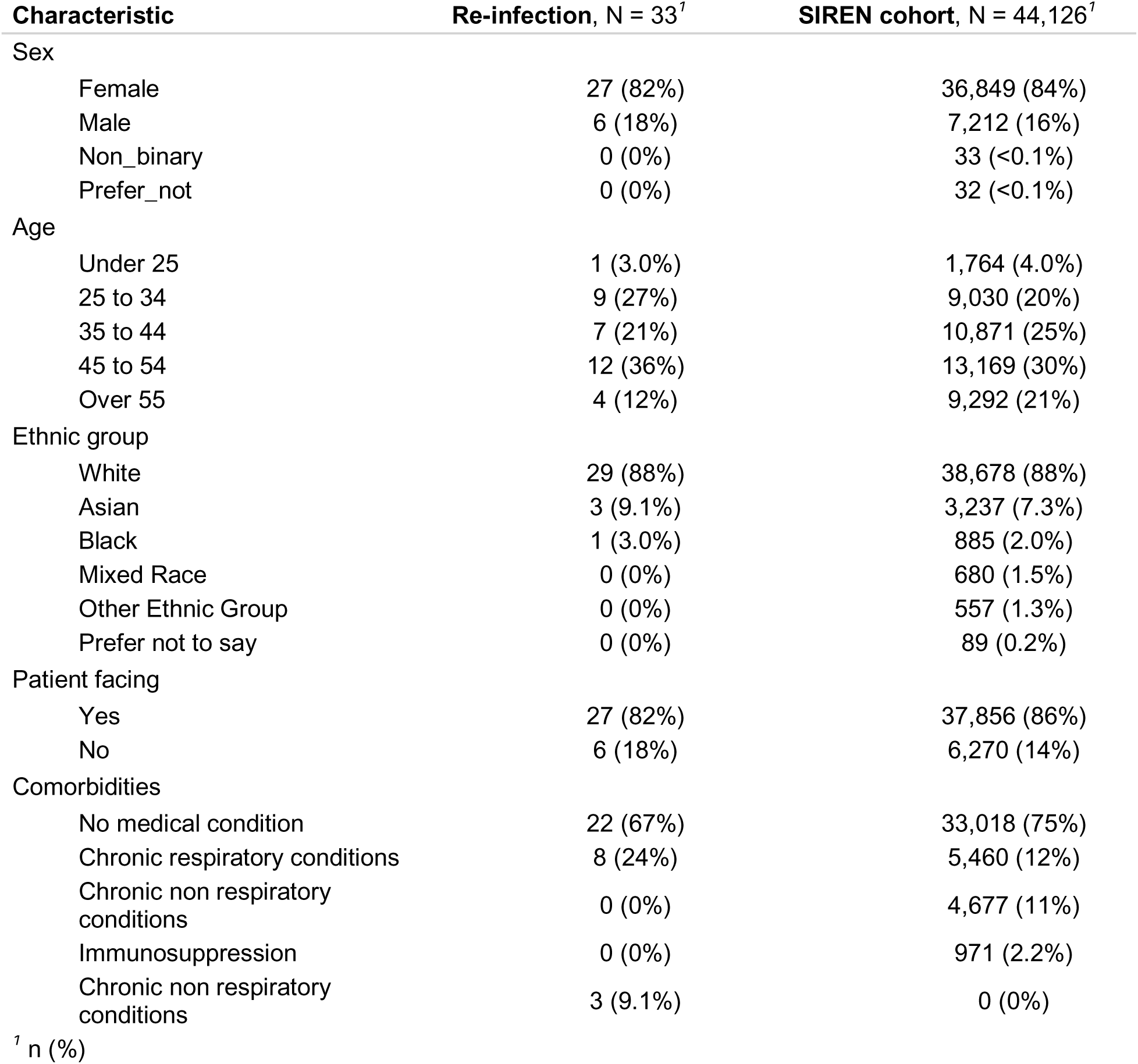
Demographics of the re-infection study compared to the whole of the SIREN study. The individuals selected for this study (n=33) are also included in the whole SIREN cohort (n=44,126).

Reinfections occurred between 20^th^ December 2021 until 21^st^ June 2022; 27% (9/33) of participants were reinfected with BA.1 and 73% (24/33) with BA.2. A comparison on demographics between participants with BA.1 and BA.2 reinfections is shown in Supplementary Table 2.

The median time between first infection and reinfections was 382 days (236,424). Regarding vaccine manufacturers, 90% (30/33) of participants received BNT162b2 mRNA for their primary course (first two doses) and 10% (3/33) received ChAdOx adenoviral; as for boosters (V3), 89% (29/33) received BNT162b2 mRNA and 11% (4/33) received Moderna mRNA. The median time between third vaccine dose and reinfection was 140 days (101, 165).

### Dynamics of antibody levels and nAbs titres post-V3 by variant of first infection

We compared at peak response and decay rates by primary infection variant. For peak responses at one-week post-V3, participants with Delta first infection had higher inferred LV-N titres than those with Alpha first infection for LV-N Ancestral complete neutralisationvs 446, p=0.002), LV-N Alpha (complete neutralisation vs 624, p=0.01) and LV-N Delta (complete neutralisation vs 552, p=0.006). Those with Delta first infection also had considerably higher LV-Omicron BA.1 titres when compared to individuals with Alpha first infection (1154 vs 436, p=0.05). No statistical differences were observed for anti-S (62861 vs 33587 BAU/mL, p=0.31) and LV-Omicron BA.2 (1232 vs 770, p=0.36) by variant of first infection (Figure 2). Time since first infection was not significantly associated with the values of titres and did not affect the estimates of the other parameters of the model.

**Figure 2.**
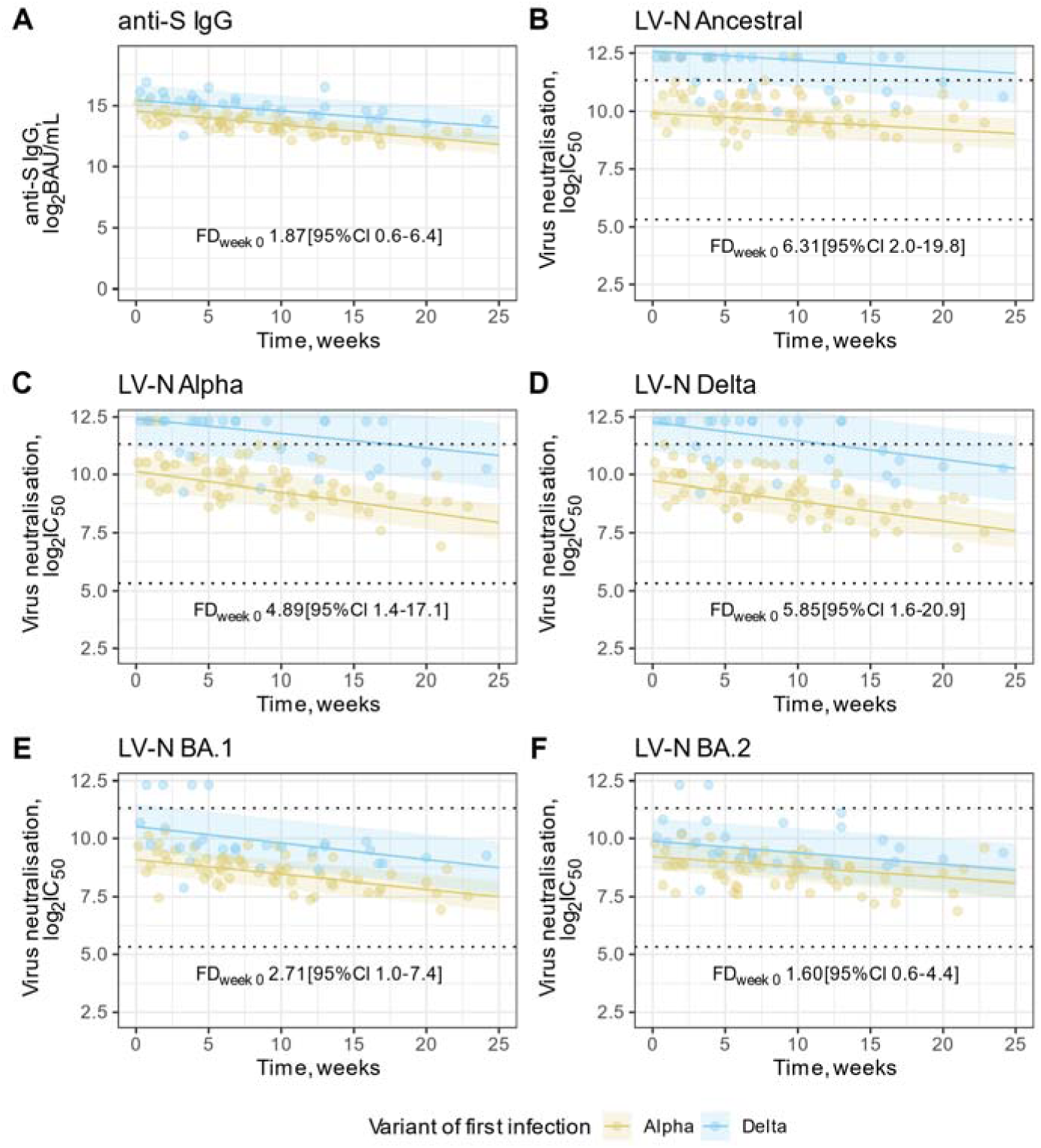
Estimated trajectories for anti-S and neutralising antibodies by variant of first infection. **(A)** anti-S IgG plotted against time since one week after each individual’s third vaccine dose (time 0 = V3+7d) for participants with an Alpha or Delta first infection. **(B-F)** live virus neutralisation (LV-N) titres plotted against time since each individual’s third vaccine dose (V3) for participants with an Alpha or Delta first infection. Ancestral, Alpha, Delta, Omicron BA.1 and Omicron BA.2 are shown in B to F, respectively. Live virus neutralisation titres are reported as the reciprocal of the dilution at which 50% inhibition of viral entry occurred (IC_50_). In (A-F), all serological data are log2-transformed and a population-level fit line and confidence intervals are shown for Alpha or Delta first infections. Fold differences (FD) at week 0 of the model fits (V3+7d) between Alpha and Delta first infections are reported on a linear scale. In (B-F), dashed horizontal lines indicate the upper and lower bounds of the quantitative range.

When analysing decay rates, we found no statistical difference by variant of first infection for all antibody responses measured (p=0.27 for anti-S, larger for nAb) (Figure 2). When fitting a model without the interaction term, estimated overall decay rates per week were: anti-S 6.9% (95% CI 5.8% to 8.3%); LV-N Ancestral 2.5% (CI 1.2% to 3.7%); LV-N Alpha 5.5%, (CI 4.0% to 6.9%), LV-N Delta 5.7%, (CI 4.3% to 7.1%), LV-N Omicron BA.1 4.5%, (CI 3.3% to 5.6%) and LV-N Omicron BA.2 3.2% (CI 1.8% to 4.6%).

### Comparing antibody response at the time of BA.1 infection by variant of first infection

We compared antibody responses by variant of first infection at the time of BA.1 infections. All 33 participants were included in this analysis. Participants reinfected with BA.1 had their reinfection at a median time of 13 weeks (11.1, 15.3) after V3. We inferred the titres of all participants 12 weeks after V3 and compared them by variant of first infection (Alpha or Delta).

Anti-S levels and nAb titres against all variants were higher in those with Delta first infection compared to Alpha first infection: anti-S 30458 vs 8939 BAU/mL (p=0.002), LV-N Ancestral 2234 vs 985 (p < 0.0001), LV-N Alpha 2043 vs 710 (p<0.0001), LV-N Delta 1920 vs 511 (p<0.0001), LV-N Omicron BA.1 746 vs 32 (p=0.001) and LV-N Omicron BA.2 861 vs 409 (p=0.0009) (Figure 3).

**Figure 3.**
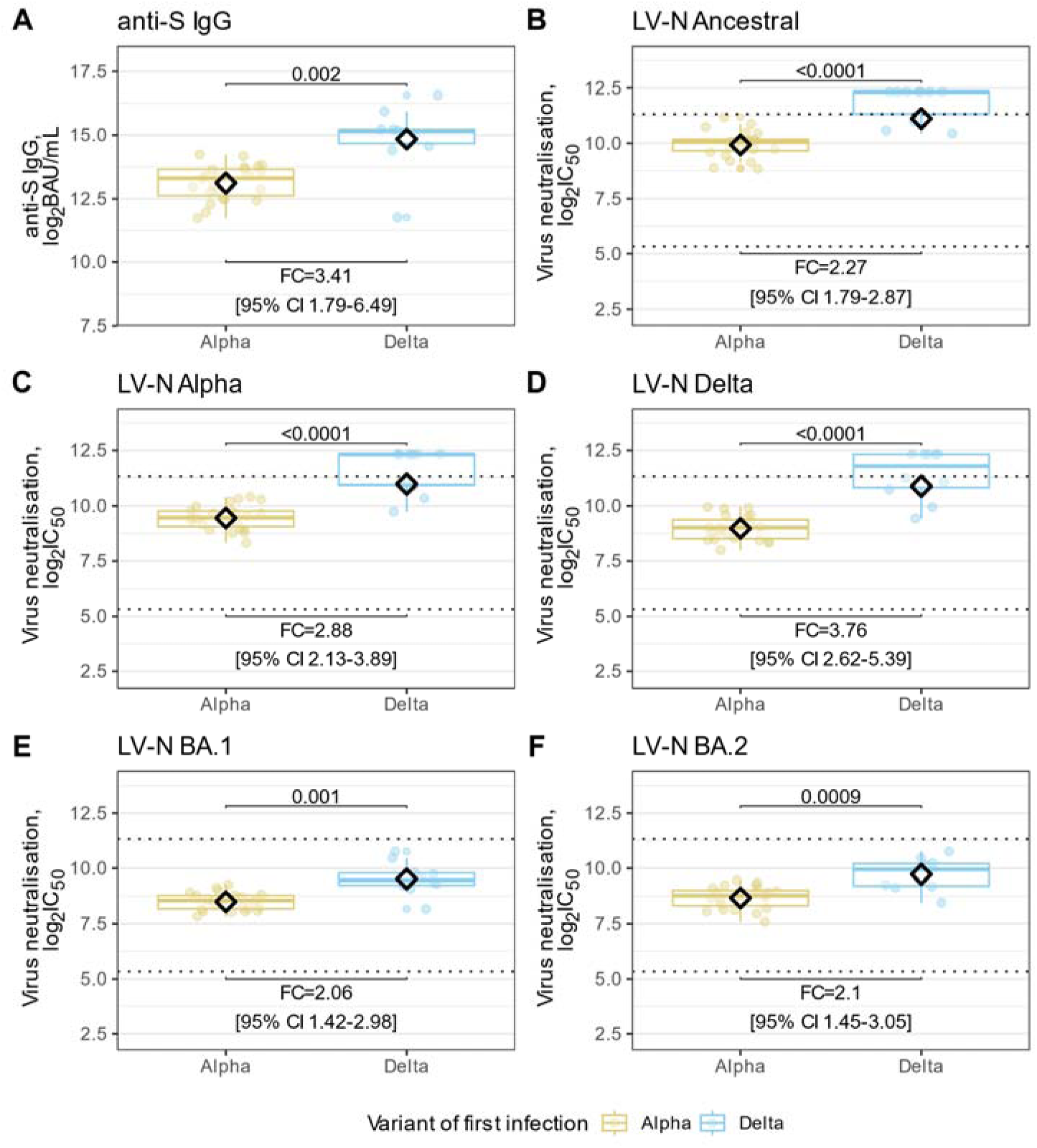
Anti-S levels and neutralising antibody titres inferred at the time of BA.1 infection, by variant of first infection. **(A)** anti-S IgG modelled at the time of BA.1 infections plotted for cohorts of Alpha or Delta first infection. **(B-F)** live virus neutralisation (LV-N) titres modelled at the time of BA.1 infections for participants plotted for cohorts of Alpha or Delta first infection. Ancestral, Alpha, Delta, Omicron BA.1 and Omicron BA.2 are shown in B to F, respectively. Live virus neutralisation titres are reported as the reciprocal of the dilution at which 50% inhibition of viral entry occurred (IC_50_). In (A-F), all serological data are log2-transformed and groups were compared. Fold differences between Alpha and Delta first infections are reported on a linear scale. Boxplots shown the interquartile range and medians, with whiskers extending to the lesser of the furthest datapoint, or 1.5x interquartile range. Diamonds indicate the geometric means. In (B-F), dashed horizontal lines indicate the upper and lower bounds of the quantitative range.

### Comparing antibody response of BA.2-infected participants by first infection variant

We compared antibody responses by variant of first infection from individuals reinfected with BA.2, 7 days before their BA.2 infection. Those with Delta first infection had higher inferred anti-S levels and nAb titres against all variants than compared to individuals with Alpha first infection: anti-S 16591 vs 5418 BAU/mL (p=0.01); LV-N Ancestral 1875 vs 826 (p=0.0001); LV-N Alpha 1875 vs 826 (p<0.0001), LV-N-Delta 1192 vs 324 (p<0.0001), LV-N Omicron BA.1 477 vs 252 (p=0.004) and LV-N Omicron BA.2 544 vs 288 (p=0.001) (Figure 4).

**Figure 4:**
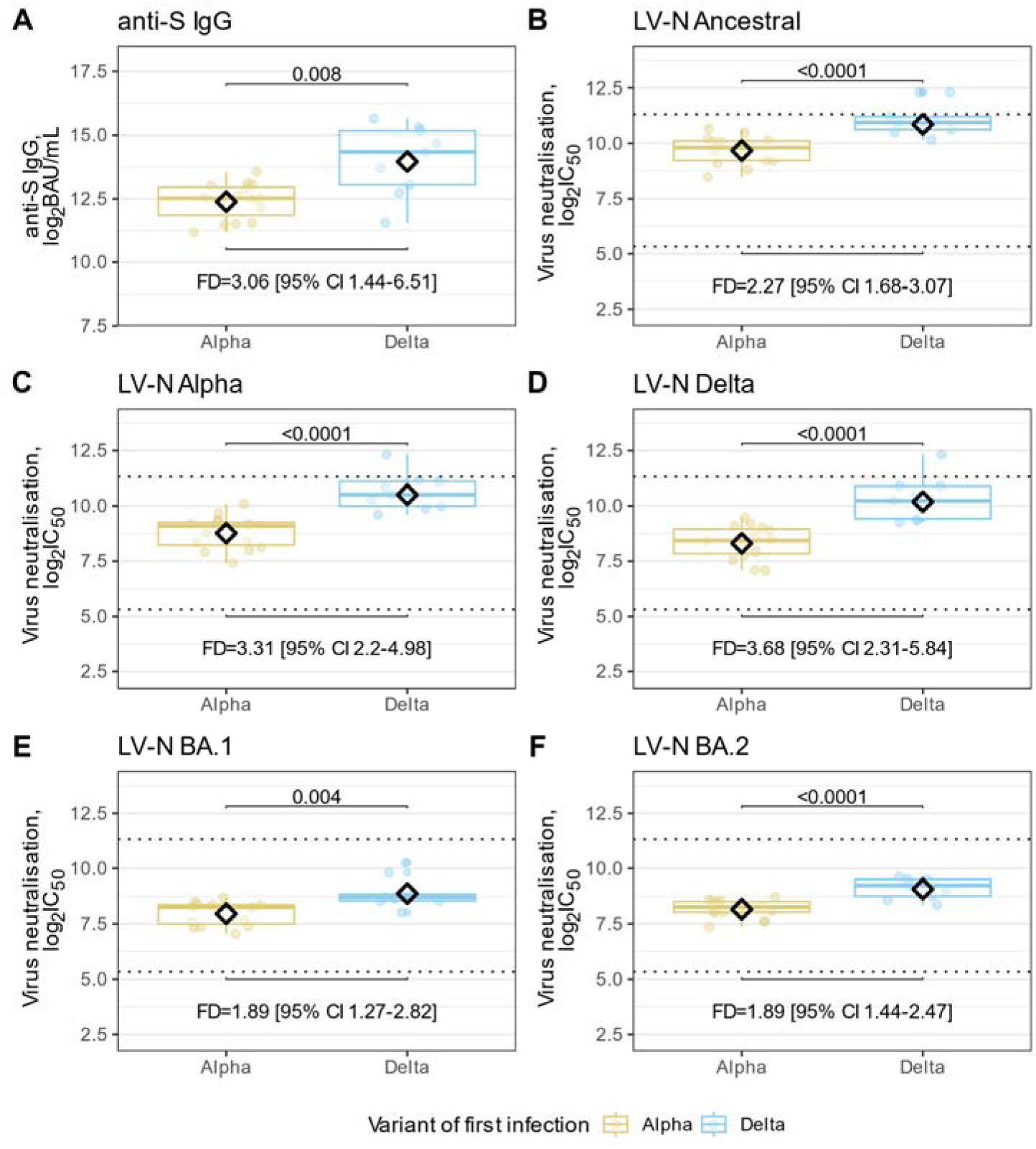
Anti-S levels and neutralising antibody titres from BA.2 reinfected participants inferred at the time of BA.2 infection, by variant of first infection. **(A)** anti-S IgG predicted at the time of BA.2 infections plotted for cohorts of Alpha or Delta first infection. **(B-F)** live virus neutralisation (LV-N) titres predicted at the time of BA.2 infections for participants plotted for cohorts of Alpha or Delta first infection. Ancestral, Alpha, Delta, Omicron BA.1 and Omicron BA.2 are shown in B to F, respectively. Live virus neutralisation titres are reported as the reciprocal of the dilution at which 50% inhibition of viral entry occurred (IC_50_). In (A-F), all serological data are log2-transformed and groups were compared. Fold differences between Alpha and Delta first infections are reported on a linear scale. Boxplots shown the interquartile range and medians, with whiskers extending to the lesser of the furthest datapoint, or 1.5x interquartile range. Diamonds indicate the geometric means. In (B-F), dashed horizontal lines indicate the upper and lower bounds of the quantitative range.

### Comparing neutralisation responses among those with Alpha or Delta first infection

To assess the potential impact of Alpha and Delta first infection in individual’s neutralisation responses following booster vaccination, we compared the neutralising antibody titres against different variants among those with Alpha (n=23) and Delta (n=10) first infection (separately). We inferred titres at 12 weeks after V3 to all 33 participants.

LV-N Ancestral titres were higher compared to titres against other variants, regardless of variant of first infection. Those with Alpha first infections had higher LV-N titres against Alpha than to other subsequent variants. For those with Delta first infections, we have still observed higher LV-N Alpha titres when compared to LV-N Delta, LV-N Omicron BA.1 and LV-N Omicron BA.2 (Figure 5).

**Figure 5:**
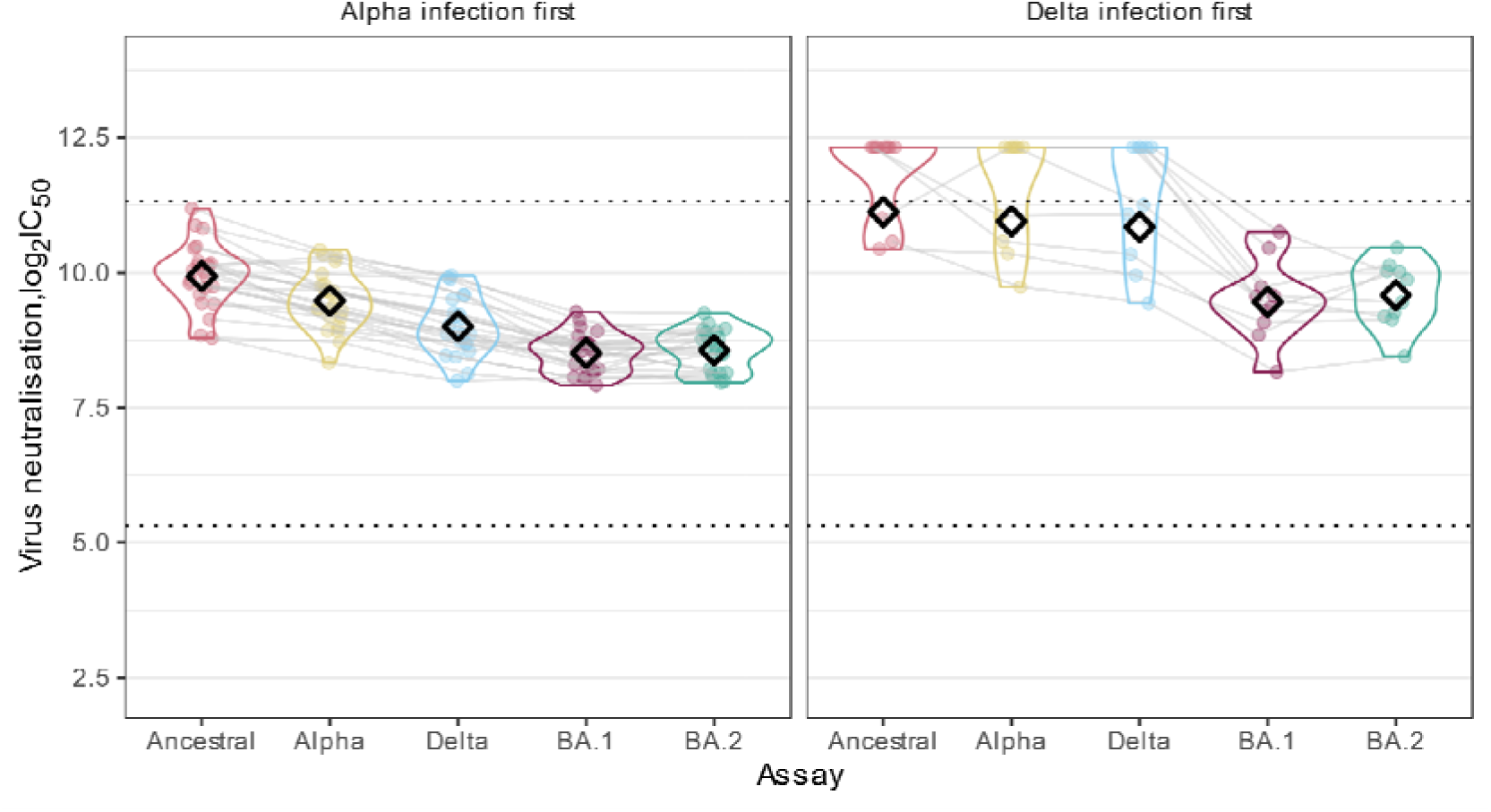
Neutralising antibody titres by variant of first infection at 12 weeks after third vaccine doses. Live virus neutralisation (LV-N) titres predicted at 12 weeks after third vaccine doses (V3), for participants with Alpha or Delta first infection. Ancestral, Alpha, Delta and Omicron BA.1 and BA.2 are shown. Live virus neutralisation titres are plotted as the log2-transformed reciprocal of the dilution at which 50% inhibition of viral entry occurred (IC_50_). Individual participants are joined by a grey line and geometric means are shown by black diamonds. Dashed horizontal lines indicate the upper and lower bounds of the quantitative range.

## DISCUSSION

Despite a large proportion of the population worldwide being previously exposed to SARS-CoV-2 infection and multiple COVID-19 vaccinations, ensuring protection against SARS-CoV-2 emerging variants remains a challenge in the face of ongoing virus evolution. Understanding the impact of first infection variant, and how it confers protection to subsequent variants, is an important topic given increasingly heterogeneous hybrid immunity. Our study with extensive longitudinal serological data paired with frequent PCR testing is well placed to investigate the impact of first infection variant in protection against Omicron (BA.1 and BA.2) infections.

We found that participants first infected with Delta variant had higher neutralising antibody titres, immediately after V3, against Ancestral, Alpha and Delta variants than those with Alpha first infection. This trend continued pre-BA.1 infection and pre-BA.2 infection. Higher antibody titres for those with Delta compared to Alpha first infection could be related to different immunogenic properties of this variant, which may lead to an enhanced inflammatory response and disease severity. (23–25) Although our study was not designed to assess disease severity, none of the participants selected for this analysis reported a hospital admission related to their first SARS-CoV-2 infection episode.

Our results suggested that a previous Delta infection conferred more protection against Omicron variants than Alpha infection, even when controlling for time since first infection. This is aligned with previous studies, which demonstrated lower protection against Omicron reinfection in those infected before Delta variant emergence when compared to those with previous Delta infection. (26, 27) It has also been demonstrated that, across multiple Omicron waves, those previously infected with more recent variants had increased protection against reinfection compared to earlier variants. (28) These findings are supported by the fact that Delta infection is more effective in inducing high nAb titres against Omicron sub-lineages as opposed to Alpha, despite BA.1 and BA.2 being antigenically distinct. (29)

When comparing an individual’s response to different variants separately for Alpha and Delta first infections following V3, higher LV-N Ancestral titres were observed regardless of variant of first infection. Although these findings provide no evidence of immunological imprinting by variant of first infection, they support the immune imprinting effect of their first antigenic exposure (Ancestral SARS-CoV-2 virus from vaccination), given most participants (79%) had received at least one vaccine dose before their first infection. Consequently, this demonstrates the immune-escape potential of early Omicron variants, with reduced cross-reactivity with predecessors’ variants. Other cohorts of individuals with different vaccination and infection records, later infected with Omicron, demonstrated similar patterns of antibody response. (30, 31) This is particularly relevant for vaccine development and supports the strategy of updating SARS-CoV-2 booster immunisations with more contemporary antigens to enhance individual’s neutralisation activity against antigenically distinct variants. (32)

Our study strengths include the frequent testing and large availability of samples, allowing us to assess the serological response in detail and provide estimates at different timepoints. Our models carefully considered different biases, such as time of first infection, that could lead to confounded results if not adjusted.

The main limitation in this study is the relatively small sample size, given we only selected participants with Omicron reinfections that had samples available at the time of data cut-off, therefore the results should be interpreted with caution. Although this may affect generalisability, our models produced results consistent with those derived from studies using larger sample sizes. In addition, there were not enough participants with distinct vaccination patterns (pre- and post-infection) to support an analysis on its impact on antibody response, which may have influenced our results. Moreover, there are many complexities intrinsic to this analysis, particularly the interconnectedness between time, variant and vaccination. Given the brisk replacement of Alpha by Delta over approximately 2 weeks in 2021, it is challenging to disentangle the effect of time since first infection and variant of first infection. Our models also considered an average time since V3 for all participants, which may have resulted in different calendar times for each participant when calculating pre-BA.1 infection titres.

In conclusion, our findings demonstrate that an Alpha or Delta primary SARS-CoV-2 infection shapes individual’s serological response against a subsequent Omicron infection.

## Ethics approval and consent to participate

The study was approved by the Berkshire Research Ethics Committee, Health Research Authority (IRAS ID 284460, REC reference 20/SC/0230) on 22 May 2020. Participants gave informed consent before joining the study.

## Authors contribution

AA, EJC, and FI designed the analysis plan and wrote the paper. AA, FI, EJC, ADO, SF, AC, RB and VH contributed to study design. SF, EJC and FI managed and finalised the dataset for analysis. EJC and FI analysed data. FI performed the statistical modelling, supervised by AC. AA did the literature search. EJC, ADO, GD, AH and MYW contributed to sample testing. All authors reviewed and approved the manuscript for publication. All authors had full access to all the data in the study and accept responsibility to submit for publication.

## Data Availability

All data produced in the present study are available upon reasonable request to the authors.

## Acknowledgements

We thank participants in the SIREN study as well as research staff at participating SIREN sites. Thanks to the UKHSA SIREN team for their dedication to the study and collaboration on this work. We would like to thank Marna Roos, Gita Mistry, Nicola Bex, Natasha Bowyer Irvine at the Francis Crick Institute. EJC is supported by a Medical Research Council Clinician Scientist Fellowship (MR/X006751/1).

