## Supplementary Table 1; Supplementary Table 2 for "Impact of first SARS-CoV-2 infection variant on serological responses against Omicron: findings from the SIREN study"

**SUPPLEMENTARY MATERIAL**

**Supplementary Tables**

| **Characteristic** | | **Alpha**, N = 23*^1^* | | **Delta**, N = 10*^1^* | |
| --- | --- | --- | --- | --- | --- |
| Sex | |  | |  | |
|  | Female | | 18 (78%) | | 9 (90%) |
|  | Male | | 5 (22%) | | 1 (10%) |
| Age | |  | |  | |
|  | Under 25 | | 1 (4.3%) | | 0 (0%) |
|  | 25 to 34 | | 7 (30%) | | 2 (20%) |
|  | 35 to 44 | | 4 (17%) | | 3 (30%) |
|  | 45 to 54 | | 10 (43%) | | 2 (20%) |
|  | Over 55 | | 1 (4.3%) | | 3 (30%) |
| Ethnic group | |  | |  | |
|  | White | | 20 (87%) | | 9 (90%) |
|  | Asian | | 3 (13%) | | 0 (0%) |
|  | Black | | 0 (0%) | | 1 (10%) |
| Patient facing | |  | |  | |
|  | Yes | | 20 (87%) | | 7 (70%) |
|  | No | | 3 (13%) | | 3 (30%) |
| Comorbidities | |  | |  | |
|  | No medical condition | | 14 (61%) | | 8 (80%) |
|  | Chronic respiratory conditions | | 6 (26%) | | 2 (20%) |
|  | Chronic non respiratory conditions | | 3 (13%) | | 0 (0%) |
| *^1^* n (%) | | | | | |

**Supplementary Table 1. Demographics of the reinfection study grouped by the variant of their first infection.**

| **Characteristic** | | **BA.1**, N = 9*^1^* | | **BA.2**, N = 24*^1^* | |
| --- | --- | --- | --- | --- | --- |
| Sex | |  | |  | |
|  | Female | | 6 (67%) | | 21 (88%) |
|  | Male | | 3 (33%) | | 3 (13%) |
| Age | |  | |  | |
|  | Under 25 | | 1 (11%) | | 0 (0%) |
|  | 25 to 34 | | 1 (11%) | | 8 (33%) |
|  | 35 to 44 | | 4 (44%) | | 3 (13%) |
|  | 45 to 54 | | 3 (33%) | | 9 (38%) |
|  | Over 55 | | 0 (0%) | | 4 (17%) |
| Ethnic group | |  | |  | |
|  | White | | 8 (89%) | | 21 (88%) |
|  | Asian | | 1 (11%) | | 2 (8.3%) |
|  | Black | | 0 (0%) | | 1 (4.2%) |
| Patient facing | |  | |  | |
|  | Yes | | 9 (100%) | | 18 (75%) |
|  | No | | 0 (0%) | | 6 (25%) |
| Comorbidities | |  | |  | |
|  | No medical condition | | 8 (89%) | | 14 (58%) |
|  | Chronic respiratory conditions | | 0 (0%) | | 8 (33%) |
|  | Chronic non respiratory conditions | | 1 (11%) | | 2 (8.3%) |
| *^1^* n (%) | | | | | |

**Supplementary Table 2. Demographics of the re-infection study grouped by variant of reinfection.**

**SIREN Study Group members**

| **Site Name** | | **First Name** | **Surname** | |
| --- | --- | --- | --- | --- |
| ALDER HEY CHILDREN'S NHS FOUNDATION TRUST | | Stephen | McWilliam | |
| ALDER HEY CHILDREN'S NHS FOUNDATION TRUST | | Beatriz | Larru | |
| ANEURIN BEVAN UNIVERSITY LHB | | John | Northfield | |
| ANEURIN BEVAN UNIVERSITY LHB | | Sean | Cutler | |
| ASHFORD AND ST PETER'S HOSPITALS NHS FOUNDATION TRUST | | Stephen | Winchester | |
| ASHFORD AND ST PETER'S HOSPITALS NHS FOUNDATION TRUST | | Samuel | Rowley | |
| BASILDON AND THURROCK UNIVERSITY HOSPITALS NHS FOUNDATION TRUST | | Anirudda | Pai | |
| BASILDON AND THURROCK UNIVERSITY HOSPITALS NHS FOUNDATION TRUST | | Stacey | Pepper | |
| BEDFORDSHIRE HOSPITALS NHS FOUNDATION TRUST | | Simantee | Guha | |
| BEDFORDSHIRE HOSPITALS NHS FOUNDATION TRUST | | Philippa | Bakker | |
| BELFAST HEALTH & SOCIAL CARE TRUST | | Clodagh | Loughrey | |
| BETSI CADWALADR UNIVERSITY LHB | | Christian | Subbe | |
| BETSI CADWALADR UNIVERSITY LHB | | Caroline | Mulvaney Jones | |
| BIRMINGHAM AND SOLIHULL MENTAL HEALTH NHS FOUNDATION TRUST | | Manny | Bagary | |
| BIRMINGHAM AND SOLIHULL MENTAL HEALTH NHS FOUNDATION TRUST | | Nadezda | Starkova | |
| BLACK COUNTRY HEALTHCARE NHS FOUNDATION TRUST | | Alison | Grant | |
| BLACK COUNTRY HEALTHCARE NHS FOUNDATION TRUST | | Rebecca | Temple-Purcell | |
| BLACKPOOL TEACHING HOSPITALS NHS FOUNDATION TRUST | | Joanne | Howard | |
| BLACKPOOL TEACHING HOSPITALS NHS FOUNDATION TRUST | | Emma | Ward | |
| BOLTON NHS FOUNDATION TRUST | | Chinari | Subudhi | |
| BOLTON NHS FOUNDATION TRUST | | Scott | Latham | |
| BRIGHTON AND SUSSEX UNIVERSITY HOSPITALS NHS TRUST | | Bethany | Davies | |
| BRIGHTON AND SUSSEX UNIVERSITY HOSPITALS NHS TRUST | | Marion | Campbell | |
| BUCKINGHAMSHIRE HEALTHCARE NHS TRUST | | Nick | Wong | |
| BUCKINGHAMSHIRE HEALTHCARE NHS TRUST | | Ruth | Penn | |
| CALDERDALE AND HUDDERSFIELD NHS FOUNDATION TRUST | | N | Wong | |
| CALDERDALE AND HUDDERSFIELD NHS FOUNDATION TRUST | | Gavin | Boyd | |
| CENTRAL AND NORTH WEST LONDON NHS FOUNDATION TRUST | | Abigail | Severn | |
| CENTRAL AND NORTH WEST LONDON NHS FOUNDATION TRUST | | Alejandro | Arenas-Pinto | |
| CHESTERFIELD ROYAL HOSPITAL NHS FOUNDATION TRUST | | Thomas | Spencer | |
| CHESTERFIELD ROYAL HOSPITAL NHS FOUNDATION TRUST | | Edward | Harris | |
| CORNWALL PARTNERSHIP NHS FOUNDATION TRUST | | Susan | Greenwood | |
| CORNWALL PARTNERSHIP NHS FOUNDATION TRUST | | Angela | Pengilly | |
| COUNTESS OF CHESTER HOSPITAL NHS FOUNDATION TRUST | | Kim | Wells | |
| COUNTESS OF CHESTER HOSPITAL NHS FOUNDATION TRUST | | Therea | Barnes | |
| CROYDON HEALTH SERVICES NHS TRUST | | C | Jones | |
| CROYDON HEALTH SERVICES NHS TRUST | | Banerjee | Subhro-Osuji | |
| CWM TAF MORGANNWG UNIVERSITY LHB | | John | Geen | |
| CWM TAF MORGANNWG UNIVERSITY LHB | | Carla | Pothecary | |
| DARTFORD AND GRAVESHAM NHS TRUST | | Tracy | Edmunds | |
| DARTFORD AND GRAVESHAM NHS TRUST | | Nihil | Chitalia | |
| DERBYSHIRE COMMUNITY HEALTH SERVICES NHS FOUNDATION TRUST | | Ben | Pearson | |
| DERBYSHIRE COMMUNITY HEALTH SERVICES NHS FOUNDATION TRUST | | Sarah | Creer | |
| DEVON PARTNERSHIP NHS TRUST | | Clare | McAdam | |
| DEVON PARTNERSHIP NHS TRUST | | Natalie | Crooks | |
| DONCASTER AND BASSETLAW TEACHING HOSPITALS NHS FOUNDATION TRUST | | Anna | Grice | |
| DONCASTER AND BASSETLAW TEACHING HOSPITALS NHS FOUNDATION TRUST | | Ken | Agwuh | |
| DORSET COUNTY HOSPITAL NHS FOUNDATION TRUST | | Jennifer | Graves | |
| DORSET HEALTHCARE UNIVERSITY NHS FOUNDATION TRUST | | Paul | Walters | |
| EAST SUFFOLK AND NORTH ESSEX NHS FOUNDATION TRUST | | Luke | Bedford | |
| EAST SUFFOLK AND NORTH ESSEX NHS FOUNDATION TRUST | | Paul | Ridley | |
| EAST SUSSEX HEALTHCARE NHS TRUST | | Anna | Cowley | |
| EAST SUSSEX HEALTHCARE NHS TRUST | | Janet | Sinclair | |
| EPSOM AND ST HELIER UNIVERSITY HOSPITALS NHS TRUST | | Helen | Johnstone | |
| EPSOM AND ST HELIER UNIVERSITY HOSPITALS NHS TRUST | | Neringa | Vilimiene | |
| FRIMLEY HEALTH NHS FOUNDATION TRUST (Frimley Park hospital) | | Manjula | Meda | |
| FRIMLEY HEALTH NHS FOUNDATION TRUST (Wexham Park hospital) | | Nicky | Barnes | |
| GEORGE ELIOT HOSPITAL NHS TRUST | | Simon | Brake | |
| GEORGE ELIOT HOSPITAL NHS TRUST | | David | Boss | |
| GLOUCESTERSHIRE HOSPITALS NHS FOUNDATION TRUST | | Chris | Ford | |
| GLOUCESTERSHIRE HOSPITALS NHS FOUNDATION TRUST | | Amanda | Selassie | |
| GOLDEN JUBILEE NATIONAL HOSPITAL | | Catherine | Sinclair | |
| GOLDEN JUBILEE NATIONAL HOSPITAL | | Val | Irvine | |
| GREAT WESTERN HOSPITALS NHS FOUNDATION TRUST | | Badrinathan | Chandrasekaran | |
| GREAT WESTERN HOSPITALS NHS FOUNDATION TRUST | | Eva | Fraile | |
| HAMPSHIRE HOSPITALS NHS FOUNDATION TRUST | | Claire | Thomas | |
| HAMPSHIRE HOSPITALS NHS FOUNDATION TRUST | | Ina | Hoad | |
| HOUNSLOW AND RICHMOND COMMUNITY HEALTHCARE NHS TRUST | | John | Omany | |
| HOUNSLOW AND RICHMOND COMMUNITY HEALTHCARE NHS TRUST | | Shekoo | Mackay | |
| HULL UNIVERSITY TEACHING HOSPITALS NHS TRUST | | Phillipa | Burns | |
| HULL UNIVERSITY TEACHING HOSPITALS NHS TRUST | | Nicholas | Easom | |
| HYWEL DDA UNIVERSITY LHB | | Tracy | Lewis | |
| IMPERIAL COLLEGE HEALTHCARE NHS TRUST | | Frances | Frances | |
| IMPERIAL COLLEGE HEALTHCARE NHS TRUST | | Graham | Pickard | |
| ISLE OF WIGHT NHS TRUST | | Emily | Macnaughton | |
| ISLE OF WIGHT NHS TRUST | | Sarah | Knight | |
| JAMES PAGET UNIVERSITY HOSPITALS NHS FOUNDATION TRUST | | Davis | Nwaka | |
| JAMES PAGET UNIVERSITY HOSPITALS NHS FOUNDATION TRUST | | Christian | Hacon | |
| KING'S COLLEGE HOSPITAL NHS FOUNDATION TRUST | | Jasmin | Islam | |
| KING'S COLLEGE HOSPITAL NHS FOUNDATION TRUST | | Ray | Chaudhuri | |
| LANCASHIRE & SOUTH CUMBRIA NHS FOUNDATION TRUST | | Robert | Shorten | |
| LANCASHIRE & SOUTH CUMBRIA NHS FOUNDATION TRUST | | Kathryn | Hollinshead | |
| LEEDS TEACHING HOSPITALS NHS TRUST | | Jacqueline | Brandon | |
| LEEDS TEACHING HOSPITALS NHS TRUST | | Kyra | Holliday | |
| LEICESTERSHIRE PARTNERSHIP NHS TRUST | | Sarah | Baillon | |
| LEICESTERSHIRE PARTNERSHIP NHS TRUST | | Samantha | Hamer | |
| LEWISHAM AND GREENWICH NHS TRUST | | Judith | Russell | |
| LEWISHAM AND GREENWICH NHS TRUST | | A | Shah | |
| LINCOLNSHIRE PARTNERSHIP NHS FOUNDATION TRUST | | Kelly | Moran | |
| LINCOLNSHIRE PARTNERSHIP NHS FOUNDATION TRUST | | Vijayendra | Waykar | |
| LIVERPOOL UNIVERSITY HOSPITALS NHS FOUNDATION TRUST | | Anu | Chawla | |
| LIVERPOOL UNIVERSITY HOSPITALS NHS FOUNDATION TRUST | | Fran | Westwell | |
| LONDON NORTH WEST UNIVERSITY HEALTHCARE NHS TRUST | | P. | Papinni | |
| LONDON NORTH WEST UNIVERSITY HEALTHCARE NHS TRUST | | Ekaterina | Watson | |
| MAIDSTONE AND TUNBRIDGE WELLS NHS TRUST | | Claire | Pegg | |
| MAIDSTONE AND TUNBRIDGE WELLS NHS TRUST | | Maureen | Williams | |
| MANCHESTER UNIVERSITY NHS FOUNDATION TRUST | | Alexander | Horsley | |
| MANCHESTER UNIVERSITY NHS FOUNDATION TRUST | | Shazaad | Ahmad | |
| MID CHESHIRE HOSPITALS NHS FOUNDATION TRUST | | Elijah | Matovu | |
| MID CHESHIRE HOSPITALS NHS FOUNDATION TRUST | | Claire | Gabriel | |
| MID ESSEX HOSPITAL SERVICES NHS TRUST | | Lauren | Sach | |
| MID ESSEX HOSPITAL SERVICES NHS TRUST | | Yvonne | Lester | |
| MID YORKSHIRE HOSPITALS NHS TRUST | | AJ. | Ashcroft | |
| MID YORKSHIRE HOSPITALS NHS TRUST | | Ismaelette | Del Rosario | |
| MOORFIELDS EYE HOSPITAL NHS FOUNDATION TRUST | | Roxanne | Crosby-Nwaobi | |
| MOORFIELDS EYE HOSPITAL NHS FOUNDATION TRUST | | Chloe | Reeks | |
| NHS BORDERS | | Joy | Dawson | |
| NHS BORDERS | | Lauren | Finlayson | |
| NHS FIFE | | Devesh | Dhasmana | |
| NHS FIFE | | Susan | Fowler | |
| NHS FORTH VALLEY | | Euan | Cameron | |
| NHS FORTH VALLEY | | Anne | Todd | |
| NHS GRAMPIAN | | Vhair | Bateman | |
| NHS GRAMPIAN | | Sally | Mavin | |
| NHS GREATER GLASGOW AND CLYDE | | Antonia | Ho | |
| NHS GREATER GLASGOW AND CLYDE | | Michael | Murphy | |
| NHS HIGHLAND | | Andrew | Gibson | |
| NHS HIGHLAND | | Alexandra | Cochrane | |
| NHS LANARKSHIRE | | Manish | Patel | |
| NHS LANARKSHIRE | | Berni | Welsh | |
| NHS LOTHIAN | | Kate | Templeton | |
| NHS LOTHIAN | | Sam | Donaldson | |
| NHS WESTERN ISLES | | Martin | Malcolm | |
| NHS WESTERN ISLES | | Beth | Smith | |
| NORFOLK AND NORWICH UNIVERSITY HOSPITALS NHS FOUNDATION TRUST | | Ngozi | Elumogo | |
| NORFOLK AND NORWICH UNIVERSITY HOSPITALS NHS FOUNDATION TRUST | | Louise | Coke | |
| NORTH CUMBRIA INTEGRATED CARE NHS FOUNDATION TRUST | | Edward | Barton | |
| NORTH CUMBRIA INTEGRATED CARE NHS FOUNDATION TRUST | | Beverley | Wilkinson | |
| NORTH MIDDLESEX UNIVERSITY HOSPITAL NHS TRUST | | Mariyam | Mirfenderesky | |
| NORTH MIDDLESEX UNIVERSITY HOSPITAL NHS TRUST | | Swati | Jain | |
| NORTH WEST ANGLIA NHS FOUNDATION TRUST | | Kanchan | Rege | |
| NORTH WEST ANGLIA NHS FOUNDATION TRUST | | Janki | Bhayani | |
| NORTHERN DEVON HEALTHCARE NHS TRUST | | Tom | Lewis | |
| NORTHERN DEVON HEALTHCARE NHS TRUST | | M | Howard | |
| NORTHERN HEALTH & SOCIAL CARE TRUST | | Elinor | Hanna | |
| NORTHERN HEALTH & SOCIAL CARE TRUST | | Frances | Johnston | |
| NORTHERN LINCOLNSHIRE AND GOOLE NHS FOUNDATION TRUST | | Jonathan | Hatton | |
| NORTHERN LINCOLNSHIRE AND GOOLE NHS FOUNDATION TRUST | | Peter | Cowling | |
| NOTTINGHAM UNIVERSITY HOSPITALS NHS TRUST | | Sarah | Brand | |
| NOTTINGHAM UNIVERSITY HOSPITALS NHS TRUST | | Jack | Squires | |
| POOLE HOSPITAL NHS FOUNDATION TRUST | | Liz | Sheridan | |
| POOLE HOSPITAL NHS FOUNDATION TRUST | | Charlotte | Barclay | |
| PORTSMOUTH HOSPITALS NHS TRUST | | Johanna | Mouland | |
| PORTSMOUTH HOSPITALS NHS TRUST | | Karen | Hudson | |
| POWYS TEACHING LHB | | Jayne | Goodwin | |
| POWYS TEACHING LHB | | Chris | Norman | |
| QUEEN VICTORIA HOSPITAL NHS FOUNDATION TRUST | | Julian | Giles | |
| ROYAL BERKSHIRE NHS FOUNDATION TRUST | | Tim | Parke | |
| ROYAL BERKSHIRE NHS FOUNDATION TRUST | | Maya | Joseph | |
| ROYAL CORNWALL HOSPITALS NHS TRUST | | Duncan | Browne | |
| ROYAL CORNWALL HOSPITALS NHS TRUST | | H | Chenoweth | |
| ROYAL DEVON AND EXETER NHS FOUNDATION TRUST | | Cressida | Auckland | |
| ROYAL DEVON AND EXETER NHS FOUNDATION TRUST | | Stephanie | Prince | |
| ROYAL FREE LONDON NHS FOUNDATION TRUST | | Alison | Rodger | |
| ROYAL FREE LONDON NHS FOUNDATION TRUST | | Tabitha | Mahungu | |
| ROYAL NATIONAL ORTHOPAEDIC HOSPITAL NHS TRUST | | Simon | Warren | |
| ROYAL NATIONAL ORTHOPAEDIC HOSPITAL NHS TRUST | | Esther | Hanison | |
| ROYAL PAPWORTH HOSPITAL NHS FOUNDATION TRUST | | Sumita | Pai | |
| ROYAL PAPWORTH HOSPITAL NHS FOUNDATION TRUST | | Allison | Doel | |
| ROYAL SURREY COUNTY HOSPITAL NHS FOUNDATION TRUST | | Chery | Marriott | |
| ROYAL SURREY COUNTY HOSPITAL NHS FOUNDATION TRUST | | Charles | Piercy | |
| ROYAL UNITED HOSPITALS BATH NHS FOUNDATION TRUST | | Debbie | Delgado | |
| ROYAL UNITED HOSPITALS BATH NHS FOUNDATION TRUST | | Julia | Vasant | |
| ROYAL UNITED HOSPITALS BATH NHS FOUNDATION TRUST | | Deborah | Howcroft | |
| ROYAL UNITED HOSPITALS BATH NHS FOUNDATION TRUST | | Sarah | Meisner | |
| SALISBURY NHS FOUNDATION TRUST | | Abby | Rand | |
| SALISBURY NHS FOUNDATION TRUST | | Catherine | Thompson | |
| SALISBURY NHS FOUNDATION TRUST | | Holly | Morgan | |
| SANDWELL AND WEST BIRMINGHAM HOSPITALS NHS TRUST | | Ash | Turner | |
| SANDWELL AND WEST BIRMINGHAM HOSPITALS NHS TRUST | | Anne | Hayes | |
| SHEFFIELD CHILDREN'S NHS FOUNDATION TRUST | | Fiona | Shackley | |
| SHEFFIELD CHILDREN'S NHS FOUNDATION TRUST | | James | Pethick | |
| SHEFFIELD TEACHING HOSPITALS NHS FOUNDATION TRUST | | Thushan | de Silva | |
| SHEFFIELD TEACHING HOSPITALS NHS FOUNDATION TRUST | | Helen | Shulver | |
| SHERWOOD FOREST HOSPITALS NHS FOUNDATION TRUST | | Lynne | Allsop | |
| SHERWOOD FOREST HOSPITALS NHS FOUNDATION TRUST | | Shrikant | Ambalkar | |
| SHREWSBURY AND TELFORD HOSPITAL NHS TRUST | | Mandy | Carnahan | |
| SHREWSBURY AND TELFORD HOSPITAL NHS TRUST | | Mandy | Beekes | |
| SHROPSHIRE COMMUNITY HEALTH NHS TRUST | | Johanne | Tomlinson | |
| SOLENT NHS TRUST | | Cathy | Price | |
| SOMERSET NHS FOUNDATION TRUST | | Justin | Pepperell | |
| SOMERSET NHS FOUNDATION TRUST | | Kate | James | |
| SOUTH EASTERN HEALTH & SOCIAL CARE | | Yuri | Protaschik | |
| SOUTH EASTERN HEALTH & SOCIAL CARE | | Susan | Regan | |
| SOUTHEND UNIVERSITY HOSPITAL NHS FOUNDATION TRUST | | John | Day | |
| SOUTHEND UNIVERSITY HOSPITAL NHS FOUNDATION TRUST | | Swapna | Kunhunny | |
| SOUTHERN HEALTH & SOCIAL CARE TRUST | | Angel | Boulos | |
| SOUTHERN HEALTH & SOCIAL CARE TRUST | | Fiona | Thompson | |
| SOUTHPORT AND ORMSKIRK HOSPITAL NHS TRUST | | Katherine | Gray | |
| SOUTHPORT AND ORMSKIRK HOSPITAL NHS TRUST | | Kerryanne | Brown | |
| ST GEORGE'S UNIVERSITY HOSPITALS NHS FOUNDATION TRUST | | Tim | Planche | |
| ST GEORGE'S UNIVERSITY HOSPITALS NHS FOUNDATION TRUST | | Angela | Houston | |
| ST HELENS AND KNOWSLEY TEACHING HOSPITALS NHS TRUST | | Rowan | Pritchard-Jones | |
| ST HELENS AND KNOWSLEY TEACHING HOSPITALS NHS TRUST | | Diane | Wycherley | |
| STOCKPORT NHS FOUNDATION TRUST | | Sharman | Harris | |
| STOCKPORT NHS FOUNDATION TRUST | | Barzo | Faris | |
| SURREY AND SUSSEX HEALTHCARE NHS TRUST | | Kofi | Nimako | |
| SURREY AND SUSSEX HEALTHCARE NHS TRUST | | Simon | Bax | |
| SWANSEA BAY UNIVERSITY LHB | | Rebeccah | Thomas | |
| SWANSEA BAY UNIVERSITY LHB | | Steve | Bain | |
| THE CLATTERBRIDGE CANCER CENTRE NHS FOUNDATION TRUST | | Sheena | Khanduri | |
| THE CLATTERBRIDGE CANCER CENTRE NHS FOUNDATION TRUST | | Nagesh | Kalakonda | |
| THE DUDLEY GROUP NHS FOUNDATION TRUST | | Helen | Ashby | |
| THE HILLINGDON HOSPITALS NHS FOUNDATION TRUST | | Ayida | Gubby | |
| THE HILLINGDON HOSPITALS NHS FOUNDATION TRUST | | Natasha | Mahabir | |
| THE NEWCASTLE UPON TYNE HOSPITALS NHS FOUNDATION TRUST | | Brendan | Payne | |
| THE NEWCASTLE UPON TYNE HOSPITALS NHS FOUNDATION TRUST | | Jayne | Harwood | |
| THE PRINCESS ALEXANDRA HOSPITAL NHS TRUST | | Kathryn | Court | |
| THE PRINCESS ALEXANDRA HOSPITAL NHS TRUST | | Nikki | White | |
| THE ROBERT JONES AND AGNES HUNT ORTHOPAEDIC HOSPITAL NHS FOUNDATION TRUST | | Ruth | Longfellow | |
| THE ROYAL BOURNEMOUTH AND CHRISTCHURCH HOSPITALS NHS FOUNDATION TRUST | | Mihye | Lee | |
| THE ROYAL WOLVERHAMPTON NHS TRUST | | Clare | Ford | |
| THE ROYAL WOLVERHAMPTON NHS TRUST | | Marie | Green | |
| TORBAY AND SOUTH DEVON NHS FOUNDATION TRUST | | Kelly | Barrett | |
| TORBAY AND SOUTH DEVON NHS FOUNDATION TRUST | | Matthew | Halkes | |
| UNITED LINCOLNSHIRE HOSPITALS NHS TRUST | | Alun | Roebuck | |
| UNIVERSITY HOSPITAL SOUTHAMPTON NHS FOUNDATION TRUST | | Eleri | Wilson-Davies | |
| UNIVERSITY HOSPITALS BRISTOL AND WESTON NHS FOUNDATION TRUST | | Rajeka | Lazarus | |
| UNIVERSITY HOSPITALS BRISTOL AND WESTON NHS FOUNDATION TRUST | | Aaran | Sinclair | |
| UNIVERSITY HOSPITALS COVENTRY AND WARWICKSHIRE NHS TRUST | | N | Aldridge | |
| UNIVERSITY HOSPITALS COVENTRY AND WARWICKSHIRE NHS TRUST | | Lisa | Berry | |
| UNIVERSITY HOSPITALS OF DERBY AND BURTON NHS FOUNDATION TRUST | | L | Berry | |
| UNIVERSITY HOSPITALS OF DERBY AND BURTON NHS FOUNDATION TRUST | | Frances | Game | |
| UNIVERSITY HOSPITALS OF LEICESTER NHS TRUST | | Christopher | Holmes | |
| UNIVERSITY HOSPITALS OF LEICESTER NHS TRUST | | Martin | Wiselka | |
| UNIVERSITY HOSPITALS OF MORECAMBE BAY NHS FOUNDATION TRUST | | Timothy | Gatheral | |
| UNIVERSITY HOSPITALS OF MORECAMBE BAY NHS FOUNDATION TRUST | | Lynda | Fothergill | |
| UNIVERSITY HOSPITALS PLYMOUTH NHS TRUST | | David | Hilton | |
| UNIVERSITY HOSPITALS PLYMOUTH NHS TRUST | | Hannah | Jory | |
| VELINDRE NHS TRUST | | Charlotte | Young | |
| VELINDRE NHS TRUST | | James | Powell | |
| WALSALL HEALTHCARE NHS TRUST | | Lisa | Richardson | |
| WALSALL HEALTHCARE NHS TRUST | | Aiden | Plant | |
| WARRINGTON AND HALTON TEACHING HOSPITALS NHS FOUNDATION TRUST | | Zaman | Qazzafi | |
| WARRINGTON AND HALTON TEACHING HOSPITALS NHS FOUNDATION TRUST | | Lisa | Ditchfield | |
| WEST SUFFOLK NHS FOUNDATION TRUST | | Margaret | Moody | |
| WEST SUFFOLK NHS FOUNDATION TRUST | | Veronica | Mendez Moro | |
| WESTERN HEALTH & SOCIAL CARE TRUST | | Tracy | Donaghy | |
| WESTERN HEALTH & SOCIAL CARE TRUST | | Maurice | O'Kane | |
| WESTERN SUSSEX HOSPITALS NHS FOUNDATION TRUST | | R | Sierra | |
| WHITTINGTON HEALTH NHS TRUST | | Chetan | Parmar | |
| WHITTINGTON HEALTH NHS TRUST | | Philippa | Kemsley | |
| WIRRAL UNIVERSITY TEACHING HOSPITAL NHS FOUNDATION TRUST | | David | Harvey | |
| WIRRAL UNIVERSITY TEACHING HOSPITAL NHS FOUNDATION TRUST | | Y | Huang | |
| WYE VALLEY NHS TRUST | | Lisa | Robinson | |
| YEOVIL DISTRICT HOSPITAL NHS FOUNDATION TRUST | | Sarah | Board | |
| YEOVIL DISTRICT HOSPITAL NHS FOUNDATION TRUST | | Andrew | Broadley | |
| YORK TEACHING HOSPITAL NHS FOUNDATION TRUST | | Claire | Brookes | |
| YORK TEACHING HOSPITAL NHS FOUNDATION TRUST | | Neil | Todd | |
| **SIREN Associated Studies and collaborators** | **First name** | | | **Surname** |
| Protective Immunity from T cells to Covid-19 in Health workers (PITCH) | Susanna | | | Dunachie |
| Protective Immunity from T cells to Covid-19 in Health workers (PITCH) | Paul | | | Klenerman |
| Protective Immunity from T cells to Covid-19 in Health workers (PITCH) | Chris | | | Duncan |
| Protective Immunity from T cells to Covid-19 in Health workers (PITCH) | Lance | | | Turtle |
| Protective Immunity from T cells to Covid-19 in Health workers (PITCH) | Alex | | | Richter |
| Protective Immunity from T cells to Covid-19 in Health workers (PITCH) | Thushan | | | De Silva |
| Protective Immunity from T cells to Covid-19 in Health workers (PITCH) | Eleanor | | | Barnes |
| Protective Immunity from T cells to Covid-19 in Health workers (PITCH) | Daniel | | | Wootton |
| Protective Immunity from T cells to Covid-19 in Health workers (PITCH) | Christopher | | | Duncan |
| Protective Immunity from T cells to Covid-19 in Health workers (PITCH) | Rebecca | | | Payne |
| The Humoral Immune Correlates for COVID-19 (HICC) consortium | Jonathan | | | Heeney |
| The Humoral Immune Correlates for COVID-19 (HICC) consortium | Helen | | | Baxendale |
| The Humoral Immune Correlates for COVID-19 (HICC) consortium | Javier | | | Castillo-Olivares |
| The Francis Crick Institute | Rupert | | | Beale |
| The Francis Crick Institute | Edward | | | Carr |
| The Francis Crick Institute | Mary | | | Wu |
| World Influenza Centre, The Francis Crick Institute | Nicola | | | Lewis |
| World Influenza Centre, The Francis Crick Institute | Ruth | | | Harvey |
| Genotype2Phenotype (G2P) | Wendy | | | Barclay |
| Genotype2Phenotype (G2P) | Maya | | | Moshe |
| Genotype2Phenotype (G2P) | Massimo | | | Palmarini |
| Genotype2Phenotype (G2P) | Brian | | | Willett |
| GenOMICC | John Kenneth | | | Baillie |
| British Society for Immunology | Jennie | | | Evans |
| British Society for Immunology | Erika | | | Aquino |
| Wellcome Sanger Institute | Ewan | | | Harrison |
| Wellcome Sanger Institute | Katie | | | Bell |
| Wellcome Sanger Institute | Ya-Lin | | | Huang |
| Wellcome Sanger Institute | Marissa | | | Knoll |
| **Public Health Agencies** | **First name** | | | **Surname** |
| UK Health Security Agency | Susan | | | Hopkins |
| UK Health Security Agency | Victoria | | | Hall |
| UK Health Security Agency | Jasmin | | | Islam |
| UK Health Security Agency | Ana | | | Atti |
| UK Health Security Agency | Omoyeni | | | Adebiyi |
| UK Health Security Agency | Nick | | | Andrews |
| UK Health Security Agency | Hannah | | | Emmett |
| UK Health Security Agency | Jonathan | | | Broad |
| UK Health Security Agency | Nish | | | Kapirial |
| UK Health Security Agency | Simone | | | Dyer |
| UK Health Security Agency | Sophie | | | Russell |
| UK Health Security Agency | Colin | | | Brown |
| UK Health Security Agency | Joanna | | | Conneely |
| UK Health Security Agency | Paul | | | Conneely |
| UK Health Security Agency | Sarah | | | Foulkes |
| UK Health Security Agency | Nabila | | | Fowles-Gutierrez |
| UK Health Security Agency | Nipunadi | | | Hettiarachchi |
| UK Health Security Agency | Jameel | | | Khawam |
| UK Health Security Agency | Edward | | | Monk |
| UK Health Security Agency | Katie | | | Munro |
| UK Health Security Agency | Andrew | | | Taylor-Kerr |
| UK Health Security Agency | Jean | | | Timeyin |
| UK Health Security Agency | Edgar | | | Wellington |
| UK Health Security Agency | Angela | | | Dunne |
| UK Health Security Agency | Dominic | | | Sparkes |
| UK Health Security Agency | Naomi | | | Platt |
| UK Health Security Agency | Anna | | | Howells |
| UK Health Security Agency | Enemona | | | Adaji |
| UK Health Security Agency | Omolola | | | Akinbami |
| UK Health Security Agency | Palak | | | Joshi |
| UK Health Security Agency | Paola | | | Barbero |
| UK Health Security Agency | Meera | | | Chand |
| UK Health Security Agency | Andre | | | Charlett |
| UK Health Security Agency | Michelle | | | Cole |
| UK Health Security Agency | Claire | | | Neill |
| UK Health Security Agency | Anne-Marie | | | O’Connell |
| UK Health Security Agency | Ferdinando | | | Insalata |
| UK Health Security Agency | Tim | | | Brooks |
| UK Health Security Agency | Maria | | | Zambon |
| UK Health Security Agency | Mary | | | Ramsay |
| UK Health Security Agency | Ayoub | | | Saei |
| UK Health Security Agency | Ezra | | | Linley |
| UK Health Security Agency | Simon | | | Tonge |
| UK Health Security Agency | Ashley | | | Otter |
| UK Health Security Agency | Silvia | | | D’Arcangelo |
| UK Health Security Agency | Cathy | | | Rowe |
| UK Health Security Agency | Amanda | | | Semper |
| UK Health Security Agency | Eileen | | | Gallagher |
| UK Health Security Agency | Robert | | | Kyffin |
| UK Health Security Agency | Kate | | | Howell |
| UK Health Security Agency | Jacqueline | | | Hewson |
| UK Health Security Agency | Iain | | | Milligan |
| UK Health Security Agency | Noshin | | | Sajedi |
| UK Health Security Agency | Davina | | | Calbraith |
| UK Health Security Agency | Caio | | | Tranquillini |
| UK Health Security Agency | Jerry | | | Ye Aung Kyaw |
| UK Health Security Agency | Sarah | | | Wallace |
| UK Health Security Agency | Yrene | | | Themistocleous |
| UK Health Security Agency | Blanche | | | Oguti |
| UK Health Security Agency | Sakib | | | Rokadiya |
| UK Health Security Agency | Hannah | | | Emmett |
| Public Health Agency Northern Ireland | Dianne | | | Corrigan |
| Public Health Agency Northern Ireland | Lisa | | | Cromey |
| Glasgow Caledonian University & Public Health Scotland | Lesley | | | Price |
| Glasgow Caledonian University & Public Health Scotland | Nicole | | | Sergenson |
| Glasgow Caledonian University & Public Health Scotland | Sally | | | Stewart |
| Glasgow Caledonian University & Public Health Scotland | Lynne | | | Haahr |
| Glasgow Caledonian University & Public Health Scotland | Desy | | | Nuryunarsih |
| Glasgow Caledonian University | Annelysse | | | Jorgenson |
| Glasgow Caledonian University | Ayodeji | | | Matuluko |
| Glasgow Caledonian University | Melanie | | | Dembinsky |
| Glasgow Caledonian University | Desmond | | | Areghan |
| Glasgow Caledonian University | Alexander | | | Olaoye |
| Public Health Scotland | Josie | | | Evans |
| Public Health Scotland | Jennifer | | | Bishop |
| Public Health Scotland | Jennifer | | | Weir |
| Public Health Scotland | Laura | | | Dobbie |
| Public Health Scotland | Andrew | | | Telfer |
| Public Health Scotland | David | | | Goldberg |
| University of St Andrews | David | | | Crossman |
| Public Health Scotland | Caitlin | | | Plank |
| Public Health Scotland | Laura | | | Naismith |
| Public Health Wales | Ellen | | | De Lacy |
| Public Health Wales | Guy | | | Stevens |
| Public Health Wales | Susannah | | | Froude |
| Public Health Wales | Linda | | | Tyson |
| Health and Care Research Wales | Yvette | | | Ellis |
| Health and Care Research Wales | Chris | | | Norman |
